# Cost-Effectiveness of Infant and Maternal RSV Immunization Strategies, in British Columbia, Canada

**DOI:** 10.1101/2025.06.27.25330432

**Authors:** Javad Taleshi, Marina Viñeta Paramo, Allison Watts, Mark Chilvers, Jeffrey Man Hay Wong, Jolanta Piszczek, Lea Separovic, Jia Hu, Danuta Skowronski, Pascal M. Lavoie, Hind Sbihi

## Abstract

**Background:** Respiratory syncytial virus (RSV) is a leading cause of lower respiratory tract infections in young children and results in significant healthcare burden and costs. To reduce the impact of RSV in this population, the monoclonal antibody palivizumab has historically been used. Recently, new preventive options have become available, including a longer-acting monoclonal antibody (nirsevimab) and a maternal vaccine (RSVpreF).

**Methods:** We developed a discrete-event simulation model using epidemiological and cost data from British Columbia, Canada, and published efficacy estimates. The model simulated a cohort of 100,000 newborns and followed them up to 24 months. We conducted the analysis from a healthcare system perspective, evaluating five immunization strategies: (1) the historical palivizumab standard of care for high-risk children; (2) nirsevimab for high- and moderate-risk children; (3) in-season maternal RSVpreF vaccination combined with nirsevimab for high-risk children; (4) in-season maternal RSVpreF plus nirsevimab for high- and moderate-risk children; and (5) nirsevimab for all infants. We conducted a sequential cost-effectiveness analysis, ordering strategies by cost, excluding dominated or extendedly dominated options, and evaluating the remaining strategies stepwise. To support policy interpretation, we also performed a pairwise analysis comparing each strategy directly with the historical standard of care.

**Results:** In the sequential analysis, strategy 2 was the most cost-effective option. Strategy 4 provided additional health gains but was not cost-effective incrementally (ICER ≈ $119,000 per QALY vs strategy 2). Strategy 5 offered the greatest overall health benefits but was the least cost-effective option. When compared directly with the historical standard of care, however, strategy 4 was cost-effective (ICER ≈ $18,000 per QALY).

**Interpretation:** These findings support policy recommendations to prioritize nirsevimab for high- and moderate-risk infants as the most cost-effective strategy. Maternal RSVpreF vaccination offers added health benefits and is cost-effective compared with the historical standard of care, though not when considered incrementally.

## 1. Introduction

RSV is the most common cause of lower respiratory tract illness (LRTI) in young children worldwide [1]. The disease burden is highest in early infancy: nearly all children are infected by RSV by age two, and 1–2% of infants require hospitalization due to RSV infection in the first year of life in high-income countries [2–4]. RSV illness results in significant direct medical costs, with the estimated annual average cost of RSV-related pediatric hospitalizations in Canada reaching $66 million [5]. In the absence of an active pediatric vaccine, efforts to protect young children have relied on passive immunoprophylaxis with the monoclonal antibody palivizumab, the long-standing standard of care, given as monthly injections during RSV season to high-risk children (e.g., those born preterm, or with severe chronic lung or cardiac conditions, or immunodeficiency). While palivizumab can reduce severe RSV outcomes in high-risk children, it has notable limitations including high product and administration cost and logistical complexity of its dosing regimen.

In 2024, two new immunization products have become available in Canada for preventing severe RSV in young children: (1) nirsevimab, a long-acting monoclonal antibody given once to protect children during an entire RSV season; and (2) RSVpreF, a prefusion F protein vaccine administered to pregnant individuals, increasing RSV antibody levels in newborns to protect them for six months. In May 2024, the Canadian National Advisory Committee on Immunization (NACI) has recommended moving toward a universal, all-infant RSV immunization program in Canada, replacing palivizumab and prioritizing nirsevimab over RSVpreF for inclusion in infant programs and considering RSVpreF in pregnant individuals on a case-by-case basis only[6]. These recommendations emphasize staged implementation based on supply, cost-effectiveness, and affordability.

Previous evaluations have examined the cost-effectiveness of nirsevimab and maternal RSVpreF immunization strategies within the province of Ontario, Canada [7,8]. However, existing studies have some limitations. The probability of RSV infection in young children is influenced by both age and seasonality [9]. Furthermore, the severity of RSV infection, duration of hospitalization and risk of intensive care unit (ICU) admission also differ in children with co-morbidities [9,10], yet previous studies did not use co-morbidity-adjusted risks and did not account for the joint effect of age and seasonality in modeling RSV risk. Moreover, they relied on 2017 estimates for hospitalization and ICU costs, whereas more recent data are now available [5], providing updated insights into current healthcare expenditures.

To address these limitations, we developed a discrete-event simulation model that incorporates age- and month-specific RSV infection risks along with co-morbidity-adjusted clinical outcomes. The analysis was also informed by the most current demographic and retrospective healthcare data available from British Columbia (BC), Canada. By aligning infection risk with the timing of immunization and accounting for individual vulnerability, the model provides a more granular framework for evaluating the cost-effectiveness of RSV immunization strategies and identifying those that offer the greatest health and economic value.

## 2. Methods and data inputs

### 2.1. Simulation Model

We developed a discrete-event simulation model to track RSV infections and associated outcomes in a birth cohort of 100,000 children. The simulation began on May 1st, immediately after the typical end of the RSV season in BC, and ran for two full RSV seasons, concluding on April 30th. To account for seasonal variation in birth timing, the cohort was divided into 12 monthly sub-cohorts, each representing approximately one-twelfth of all births. Each child was followed from birth until the end of their second RSV season, enabling the model to capture exposure to RSV over two consecutive seasons. This extended time horizon was selected to reflect the potential long-term impact of immunization strategies, especially those that provide protection beyond the first year of life.

The model advanced in monthly time steps, during which individuals transitioned between health states, as illustrated in Figure 1A. All children entered the simulation in a healthy state, which denotes not infected with RSV. In any given month, a child could remain healthy, acquire an RSV infection, or—rarely—die from non-RSV causes. If infected with RSV, the child could either recover and return to the healthy state or die.

**Figure 1.**
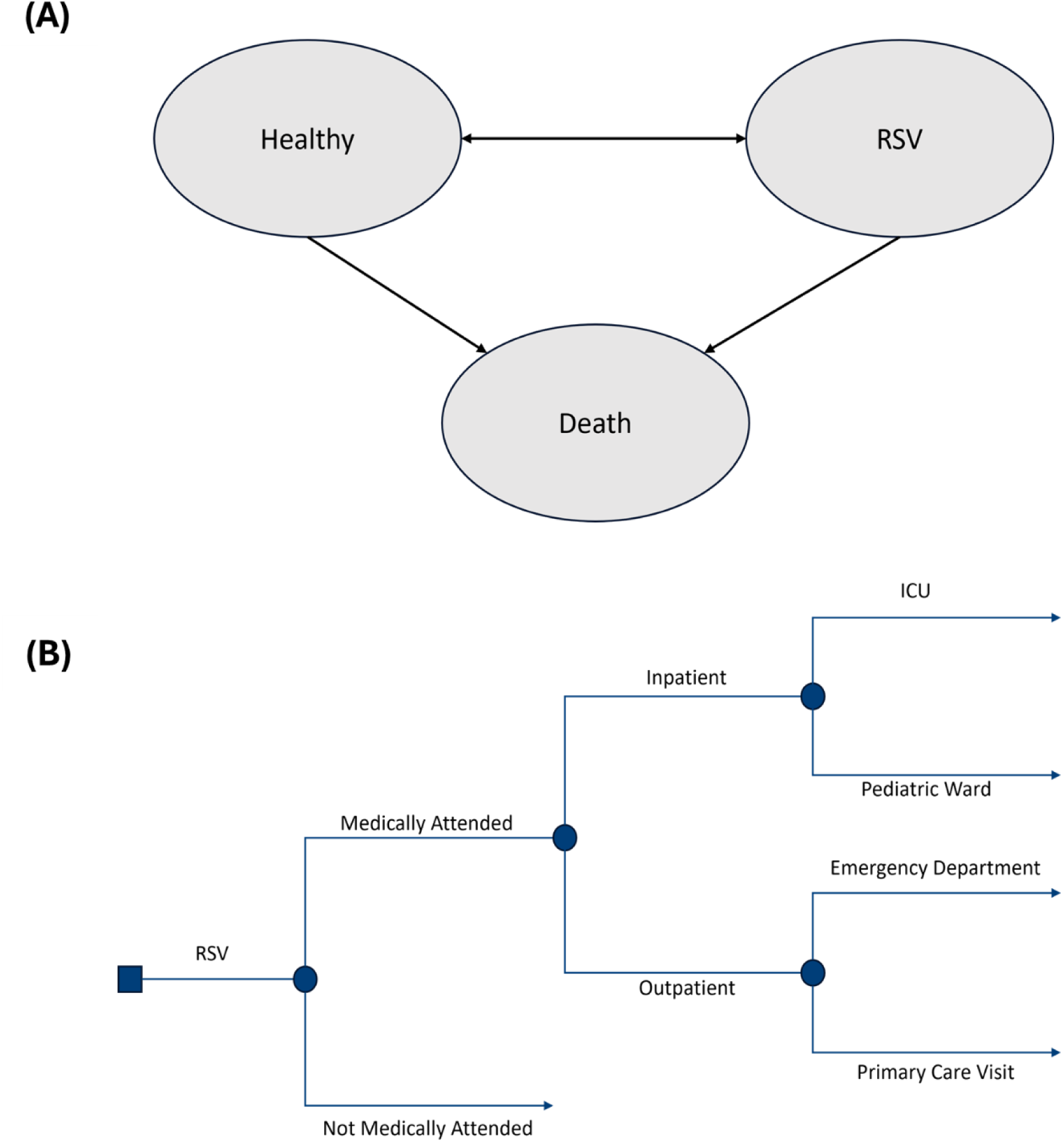
(A) Overview of health states. (B) Structure of treatment pathways for RSV infections.

In the model, an RSV infection can lead to different levels of healthcare utilization, as illustrated in Figure 1B. Children with RSV illness may receive medical care or not, and those receiving such care are further categorized into outpatient vs. inpatient management. Outpatients can be seen either in an emergency department or by a primary care provider, while inpatients are treated in hospital pediatric wards or in the ICU. Preventive interventions (monoclonal antibodies or vaccines) are assumed to reduce the probability of RSV infection and subsequent healthcare utilization, including the likelihood of severe and critical disease requiring pediatric ward and ICU admission.

### 2.2. Population data input and Hospitalization Modeling

The primary data for this analysis were drawn from a population-based cohort of children under two years of age, born between 2013 and 2023 and residing in BC. This cohort is maintained within the Provincial Health Services Authority’s (PHSA) Platform for Analytics and Data (PANDA) [11] and integrates information from provincial registries and administrative datasets, including hospitalization records and Vital Statistics [12–14]. We stratified the study cohort into three risk groups based on perinatal factors and chronic conditions, consistent with previous analyses of RSV severity during the first two seasons [9]:

- **High-risk:** Young children who would have been eligible for palivizumab under previous BC criteria (e.g., very preterm births or severe chronic medical conditions) [15]. This group represents just over 1% of children in BC, who show the highest RSV hospitalization risk.
- **Moderate-risk:** Young children born moderately preterm (28–36 weeks gestation) or those with chronic medical conditions, as we previously defined[9], and who are not classified as high-risk under the previous BC palivizumab program. Approximately 11% of children in BC fall into this group.
- **Low-risk:** The remaining young children (∼88%) who are born full-term and do not have chronic medical conditions.

Summary statistics for demographic variables used in the simulation are presented in Table 1.

**Table 1.**
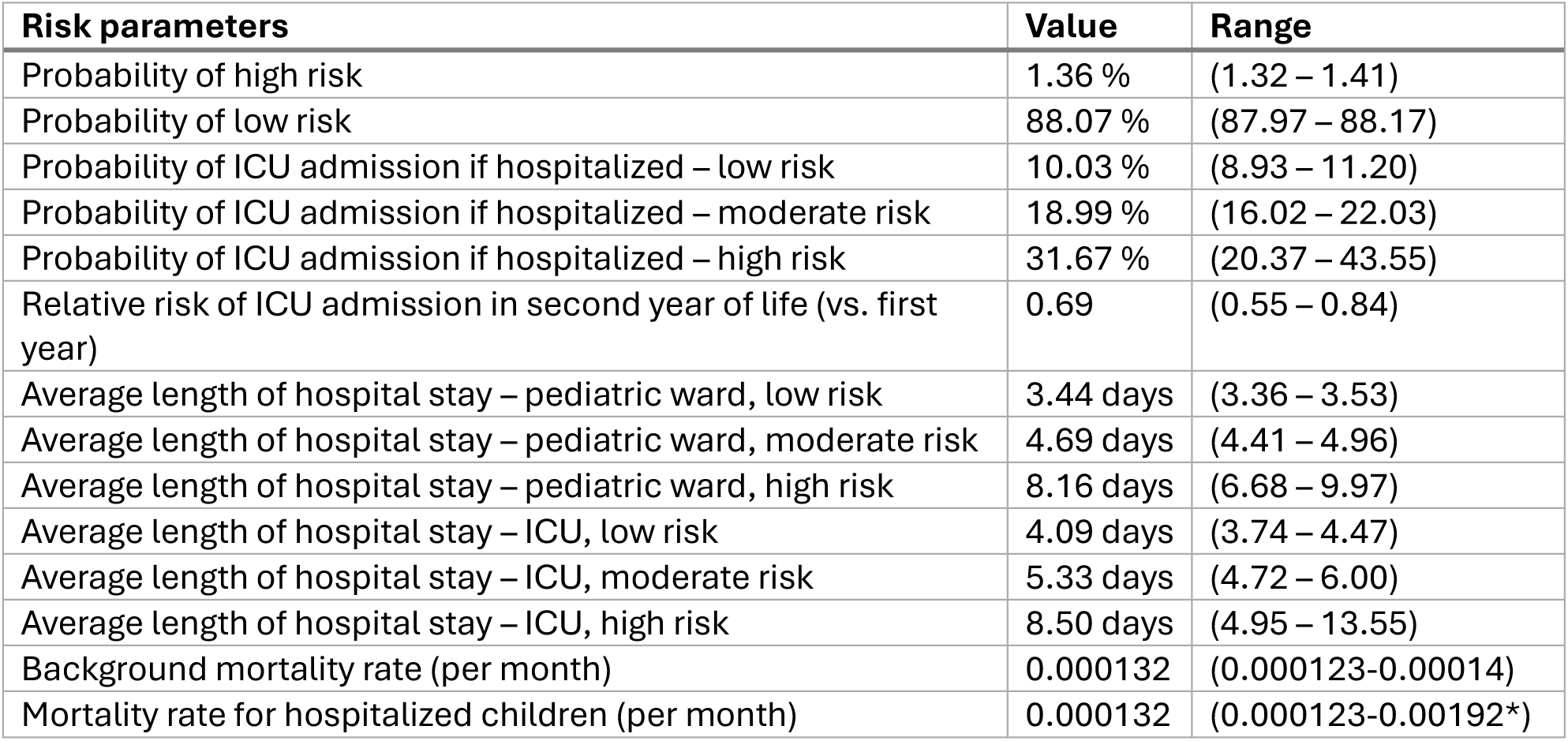
Clinical risk parameters (BC data). * Upper range value is from literature.

| **Risk parameters** | **Value** | **Range** |
| --- | --- | --- |
| Probability of high risk | 1.36 % | (1.32 – 1.41) |
| Probability of low risk | 88.07 % | (87.97 – 88.17) |
| Probability of ICU admission if hospitalized – low risk | 10.03 % | (8.93 – 11.20) |
| Probability of ICU admission if hospitalized – moderate risk | 18.99 % | (16.02 – 22.03) |
| Probability of ICU admission if hospitalized – high risk | 31.67 % | (20.37 – 43.55) |
| Relative risk of ICU admission in second year of life (vs. first year) | 0.69 | (0.55 – 0.84) |
| Average length of hospital stay – pediatric ward, low risk | 3.44 days | (3.36 – 3.53) |
| Average length of hospital stay – pediatric ward, moderate risk | 4.69 days | (4.41 – 4.96) |
| Average length of hospital stay – pediatric ward, high risk | 8.16 days | (6.68 – 9.97) |
| Average length of hospital stay – ICU, low risk | 4.09 days | (3.74 – 4.47) |
| Average length of hospital stay – ICU, moderate risk | 5.33 days | (4.72 – 6.00) |
| Average length of hospital stay – ICU, high risk | 8.50 days | (4.95 – 13.55) |
| Background mortality rate (per month) | 0.000132 | (0.000123-0.00014) |
| Mortality rate for hospitalized children (per month) | 0.000132 | (0.000123-0.00192*) |

We estimated the monthly RSV-related hospitalization rate among children under two years of age in the study cohort to define the RSV season for the microsimulation. Rates reflect the monthly averages over the 2013–2023 period, excluding data from May 1, 2020 to April 30, 2021, during which RSV activity collapsed due to public health measures implemented during the COVID-19 pandemic. As shown in Figure 2, the RSV season in BC typically spans from early November to the end of April.

**Figure 2.**
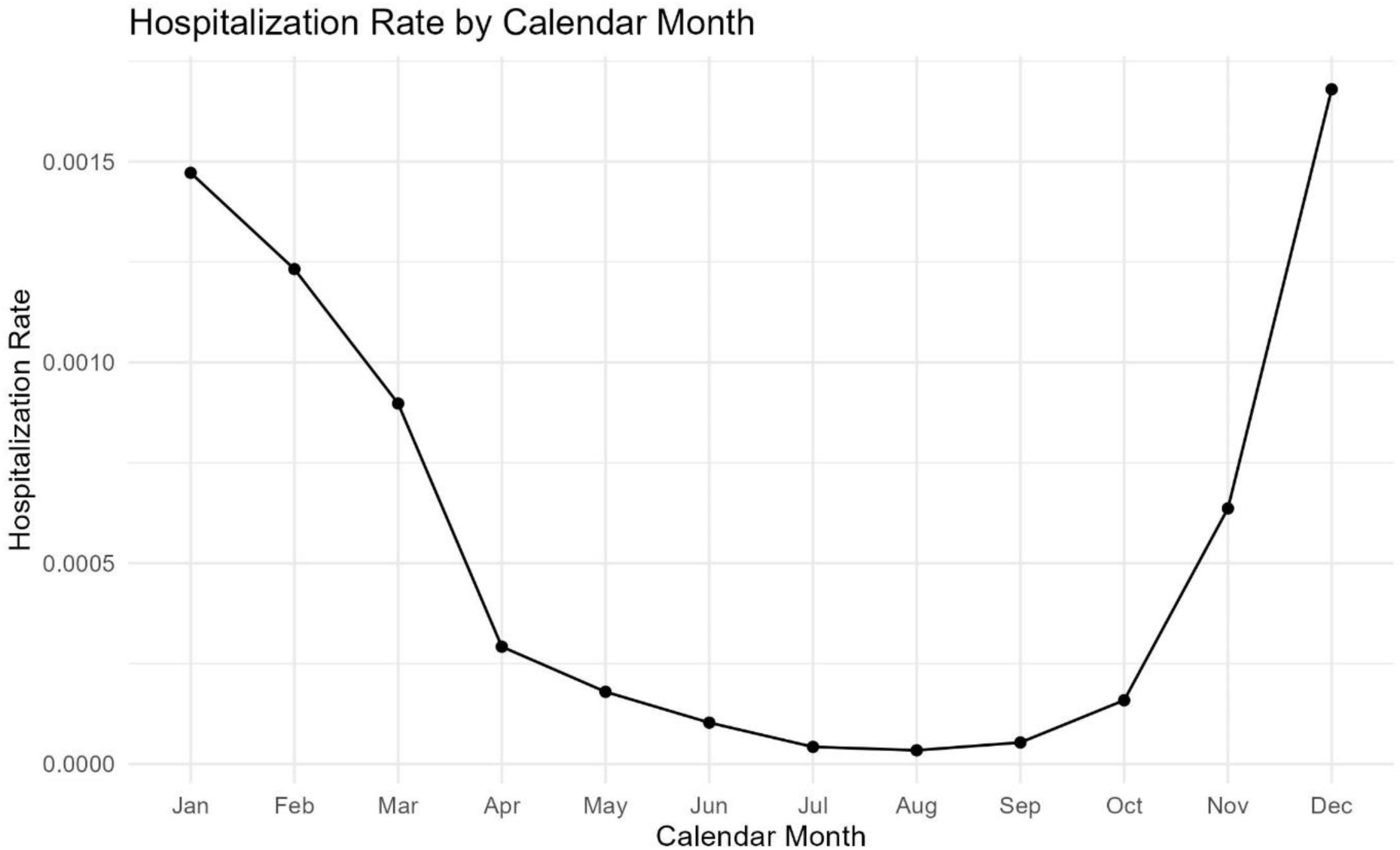
Monthly RSV hospitalization rate (per child) among children under two years of age, by calendar month of infection. Rates represent the monthly averages from 2013 to 2023.

We modeled the monthly probability of RSV hospitalization using logistic regression, incorporating age, a key determinant of RSV severity [9], palivizumab prophylaxis status, risk group, and month of infection. Model specifications and coefficients are detailed in the supplementary material (see Figure S1, Equation S1, and Table S1). The predicted probabilities showed strong agreement with observed age-specific seasonal patterns, as illustrated in Figure 3.

**Figure 3.**
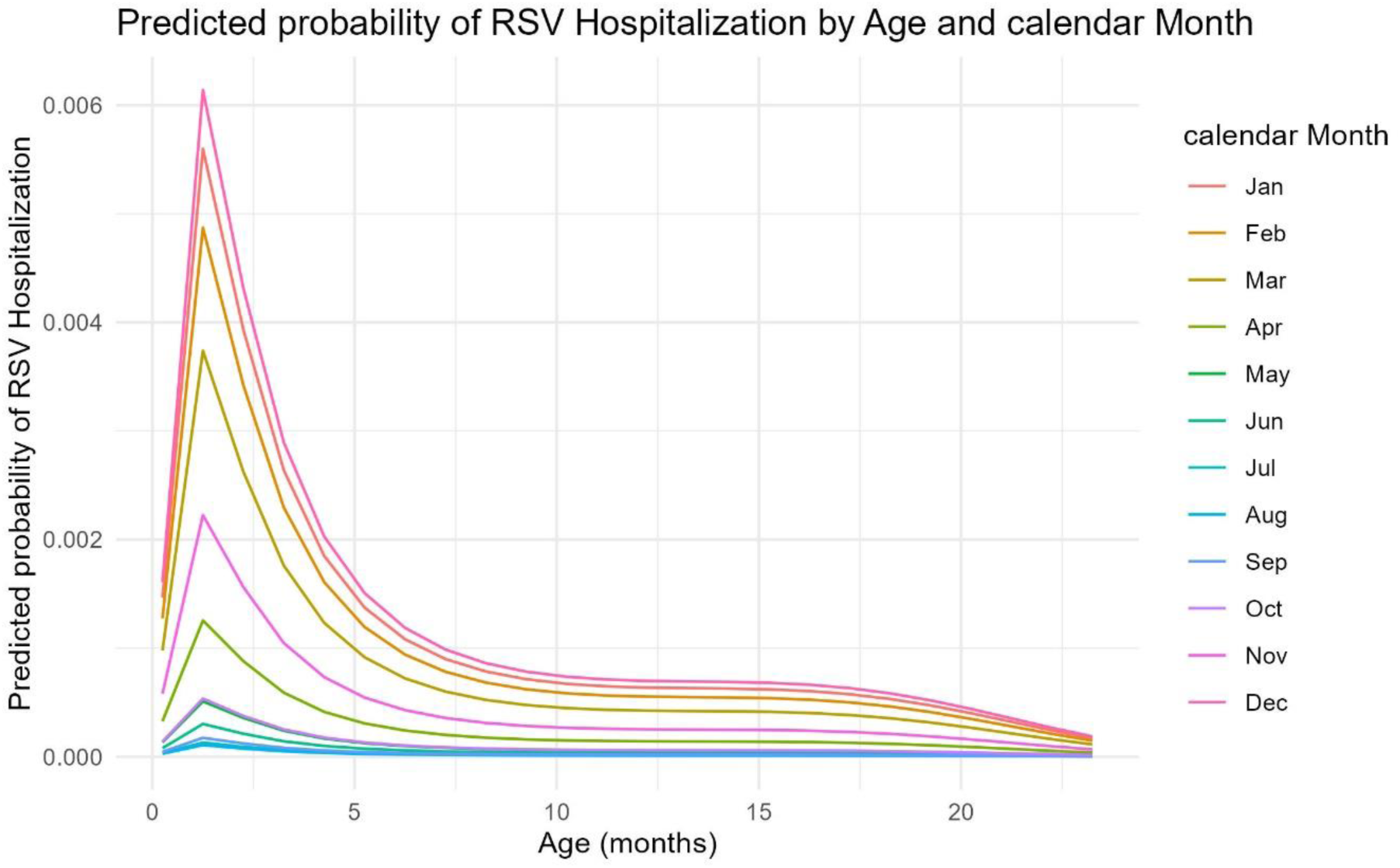
Predicted monthly probability of RSV hospitalization by age for low-risk children, without receiving palivizumab.

For high-risk children who received palivizumab prophylaxis, we adjusted the hospitalization probability based on palivizumab efficacy, reflecting the reduced observed hospitalization rates in this group.

For each risk group, we modeled the probability of ICU admission conditional on hospitalization (Table 1). We also estimated the length of stay for both pediatric ward and ICU admissions. A small monthly background mortality rate was applied, and we assumed that the mortality rate among hospitalized children was equivalent to this background rate, reflecting the very low RSV-specific mortality observed in BC [10,14]. Each death was associated with a discounted loss of 45.3 QALYs [7].

We also used published literature to obtain age-specific rates of medically attended RSV infections, which informed the distribution of RSV cases managed in outpatient settings [16]. Using these data, we estimated the proportion of RSV illnesses resulting in hospital admissions versus outpatient care by child age. These proportions enabled us to calibrate the overall probability of any medically attended RSV illness and the probability of an emergency department visit among outpatient cases.

### 2.3. Costs and Utilities

The cost-effectiveness analysis was conducted from the healthcare perspective, specifically a universal public payer system, considering only direct medical costs and utility losses experienced by children up to two years with RSV. Costs were assigned to RSV-related healthcare encounters, and immunization programs. Table 2 summarizes the cost inputs and health state utilities (QALY loss) applied in the model. All costs are in Canadian dollars (CAD).

**Table 2.** Direct medical costs and QALY losses associated with RSV-related healthcare encounters and immunization. All costs are reported in Canadian dollars (CAD). * Range defined as ±25% of the base value.

| Cost and disutility parameter | Value | Range | Reference |
| --- | --- | --- | --- |
| **Direct healthcare costs** |  |  |  |
| ICU (per day) | \$4,615 | (3461-5769) * | [5] |
| Pediatric ward (per day) | \$2,283 | (1712- 2854) * | [5] |
| Emergency department (per visit) | \$342 | (257- 428) * | [7,28] |
| Primary care (per visit) | \$ 229 | (172- 286) * | [7] |
| **Immunization costs** |  |  |  |
| RSVpreF and nirsevimab administration cost | \$15 | (0-30) | [29,30] |
| RSVpreF maternal vaccine cost per dose | \$125 | (100- 250) | Estimated |
| Nirsevimab cost per dose | \$450 | (400- 950) | Estimated |
| Palivizumab administration cost | \$150 | (0- 188*) | Estimated |
| Palivizumab cost per RSV season | \$3,600 | (2700- 4500) * | Estimated |
| **QALY loss** |  |  |  |
| QALY loss per ICU admission | 0.0245 | (0.0145–0.105) | [8] |
| QALY loss per pediatric ward admission | 0.0169 | (0.01–0.0726) | [31,32] |
| QALY loss per emergency department visit | 0.0135 | (0.008– 0.0454) | [31,32] |
| QALY loss per primary care visit | 0.00845 | (0.005–0.0454) | [31,32] |

Costs for hospital care are based on recent Canadian estimates of pediatric hospitalization costs [5]. Outpatient costs represent the estimated 30-day healthcare expenditures associated with an RSV infection not requiring hospitalization. These costs include physician visits, diagnostics, and any short-term treatments within a month of illness.

Unit prices for nirsevimab and RSVpreF are not publicly available and subject to change but we approximated their anticipated current cost as specified in Table 2 plus a $15 administration cost per dose.

Health outcomes were measured in QALYs. We assigned a small decrement in QALYs for each RSV illness, proportional to its severity, based on disutility values reported in prior cost-effectiveness analyses [8].

### 2.4. Immunization Strategies and Assumptions

The efficacy of nirsevimab and RSVpreF was drawn from phase 3 clinical trial estimates, using efficacy against severe medically attended RSV illness as a proxy for protection against ICU-level outcomes [17,18]. We modeled efficacy of both RSVpreF and nirsevimab as constant for the first five months post-immunization, followed by a linear decline to zero by ten months (see Figure S2 in the supplementary material). This waning pattern is consistent with assumptions made in prior studies [7]. Detailed information on vaccine efficacy and coverage is provided in Table S2 of the supplementary material.

Historically, all high-risk children were eligible for palivizumab prophylaxis in their first RSV season, with some also receiving it in their second season [9]. Under the evaluated strategies, nirsevimab would fully replace palivizumab prophylaxis for high-risk children, covering both their first and, as indicated by previous palivizumab eligibility, second RSV seasons. Moderate- and low-risk children, who did not receive palivizumab prophylaxis would become eligible for nirsevimab in select strategies, but only during their first RSV season.

We modeled the timing of prophylaxis to maximize protection during the RSV season. If a child was born during the RSV season (Nov–Apr), nirsevimab (or palivizumab) was administered in the birth month. If a child was born outside the season (May–Oct), prophylaxis was given at the season’s start in November. Similarly, RSVpreF maternal vaccination was assumed to be given late in pregnancy (around 32–36 weeks gestation) if the child’s birth would occur during the season. If the birth did not occur during the RSV season, RSVpreF maternal vaccination would not be given, making this an in-season only RSVpreF strategy. This seasonal approach aligns with NACI guidance and ensures stronger protection during periods of highest RSV risk.

We compared five immunization strategies in the analysis, as described below (the first being the historical standard of care). Each strategy’s target population and timing are defined based on the above criteria:

- **Palivizumab (Historic Standard of Care):** Only high-risk children born during the RSV season are eligible for palivizumab. This represented the standard of care in BC up to 2024 and serves as the referent strategy against which alternative options are compared in the pairwise analysis.
- **Nirsevimab (High & Moderate):** All high-risk and moderate-risk children are eligible for nirsevimab. Children born before the RSV season receive nirsevimab at the start of the season, while those born during the season are immunized at birth.
- **RSVpreF + Nirsevimab (High):** Maternal RSVpreF vaccination is offered to pregnant individuals with expected delivery during the RSV season. In addition, high-risk children receive nirsevimab irrespective of maternal vaccination status.
- **RSVpreF + Nirsevimab (High & Moderate):** Maternal RSVpreF vaccination is offered to pregnant individuals expected delivery during the RSV season. Additionally, all high-risk children are eligible for nirsevimab regardless of maternal vaccination status, while moderate-risk children are eligible only if they are not protected through maternal vaccination.
- **Nirsevimab (All-infant):** All children, regardless of risk status, are eligible for nirsevimab. Young children born before the RSV season are immunized at the start of the season, while those born during the season receive nirsevimab at birth.

We assumed coverage rates for these immunization programs based on emerging data (see Table S2 in the supplementary material). For maternal RSV vaccination, we assumed about 65% coverage of eligible pregnancies, reflecting uptake in a relatively new recommended maternal vaccine program [19]. For nirsevimab, we assumed 70% coverage among eligible low-risk and moderate-risk children [8], and 100% uptake among young children identified as high-risk based on observed experience with palivizumab acceptance rates.

For each strategy, the simulation was run to tally outcomes over the cohort’s first two RSV seasons. Key outcomes included the number of RSV-related medical visits (primary care visits, emergency department visits), hospitalizations (ward and ICU), total costs (healthcare costs plus immunization-related costs), total QALYs, and ICERs.

Our primary analysis used a sequential (fully incremental) cost-effectiveness framework, in which strategies were ordered by increasing cost, strictly dominated options were removed, and strategies subject to extended dominance were excluded. ICERs were then calculated relative to the next-best non-dominated alternative. This approach provides a stepwise comparison among feasible program options. To support policy decision-making in BC, we also report pairwise comparisons versus the historical standard of care, allowing policymakers to consider expansions relative to existing practice.

We applied an incremental cost-effectiveness ratio (ICER) threshold of CAD 50,000 per QALY as the willingness-to-pay benchmark. This value reflects long-standing practice in North American cost-effectiveness literature and is frequently used in Canadian analyses as an illustrative benchmark [8,20]. However, CDA has not formally adopted a single fixed ICER cutoff.

### 2.5. Input parameters for sensitivity analyses

To assess the robustness of our base-case findings, we conducted a series of sensitivity analyses using net monetary benefit (NMB) as the primary outcome. NMB is defined as:

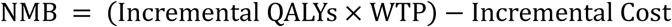

where WTP represents the willingness-to-pay threshold, set at $50,000 per QALY in this analysis. An intervention demonstrating a positive NMB is thus considered cost-effective at the specified WTP threshold.

We performed one-way sensitivity analyses by varying individual input parameters across their plausible ranges, focusing on the strategy identified as most favorable in the base-case analysis. For demographic and clinical parameters, the ranges were generally derived from 95% confidence intervals of estimates informed by provincial administrative data. An exception was the mortality rate for hospitalized children, for which the upper bound (0.00192) was based on published literature estimates [8]. For cost parameters, we varied inputs by ±25% around their base-case values unless more reliable estimates were available. Utility inputs were sourced from the literature, with ranges reflecting reported uncertainty. A complete listing of parameter ranges and sources is provided in Tables 1 and 2.

In addition to the one-way analysis, we conducted a two-way sensitivity analysis to evaluate the combined effect of varying the unit prices of nirsevimab and RSVpreF across all strategies. A broader range of prices was selected to identify the price combinations under which each strategy yields the highest net monetary benefit (NMB) relative to the alternatives.

## 3. Results

### 3.1. Health Outcomes and cost

We simulated all five immunization strategies and estimated RSV-related health outcomes for the birth cohort under each scenario. Table 3 summarizes the projected number of RSV cases requiring different levels of healthcare utilization, along with the associated costs and QALYs for each strategy.

**Table 3.** RSV-related health outcomes and costs for a simulated cohort of 100,000 infants through the end of the second RSV season, by immunization strategy. The health outcomes include ICU admissions, Pediatric Ward admissions, Emergency Department visits, Primary Care visits, and total QALYs.

| Strategies | Total Cost | Immunization Cost | ICU | PW<sup>1</sup> | ED<sup>2</sup> | PC<sup>3</sup> | Total QALY |
| --- | --- | --- | --- | --- | --- | --- | --- |
| Palivizumab (High) | \$22,401,120 | \$2,942,531 | 137 | 1044 | 7510 | 19047 | 143733 |
| Nirsevimab (High & Mod) | \$21,242,896 | \$4,119,551 | 111 | 956 | 6948 | 17504 | 143756 |
| RSVpreF + Nirsevimab (High) | \$21,869,408 | \$5,166,464 | 107 | 894 | 6867 | 17411 | 143759 |
| RSVpreF + Nirsevimab (High & Mod<sup>4</sup>) | \$23,071,289 | \$7,481,164 | 95 | 846 | 6570 | 16565 | 143772 |
| Nirsevimab (All-infant) | \$45,327,835 | \$32,799,077 | 74 | 647 | 5437 | 13347 | 143818 |

<sup>1</sup> Pediatric Ward
<sup>2</sup> Emergency Department
<sup>3</sup> Primary Care
<sup>4</sup> Moderate

The historical palivizumab strategy for high-risk children results in the highest RSV-related healthcare burden among the evaluated options. Expanding coverage with nirsevimab to include high- and moderate-risk children reduces hospitalizations and overall costs, yielding health gains at a lower total cost.

Combining maternal RSVpreF vaccination for in-season births with nirsevimab for high-risk children offers further improvements in health outcomes, with only a modest increase in total cost. Adding nirsevimab for moderate-risk children to this combined strategy reduces the disease burden even more but raises overall expenditures above the historical standard of care.

An all-infant nirsevimab strategy results in the largest health benefits, substantially lowering RSV-related disease burden. However, it also incurs significantly higher vaccination costs, making it the most expensive option evaluated.

### 3.2. Cost-Effectiveness of Immunization Program Options

Figure 4 presents the sequential cost-effectiveness analysis comparing all strategies to each other and to the historical standard of care, from the health system perspective. Strategies not on the efficient frontier are considered dominated or weakly dominated. The nirsevimab strategy for high- and moderate-risk children is cost-saving relative to the historical standard of care and lies on the efficient frontier. In contrast, the maternal RSVpreF plus high-risk nirsevimab strategy is weakly dominated and therefore excluded from the incremental sequence. The combined maternal RSVpreF plus nirsevimab (high & moderate)strategy is also on the efficient frontier, showing additional health gains at higher program costs. Incremental costs, QALYs, and ICERs for each strategy compared with the next-best non-dominated alternative are summarized in Table 4A. This framework allows stepwise comparison among feasible program options after excluding dominated and extendedly dominated strategies. The corresponding pairwise comparisons versus the historical standard of care are available in the Table 4B for policy reference.

**Figure 4.**
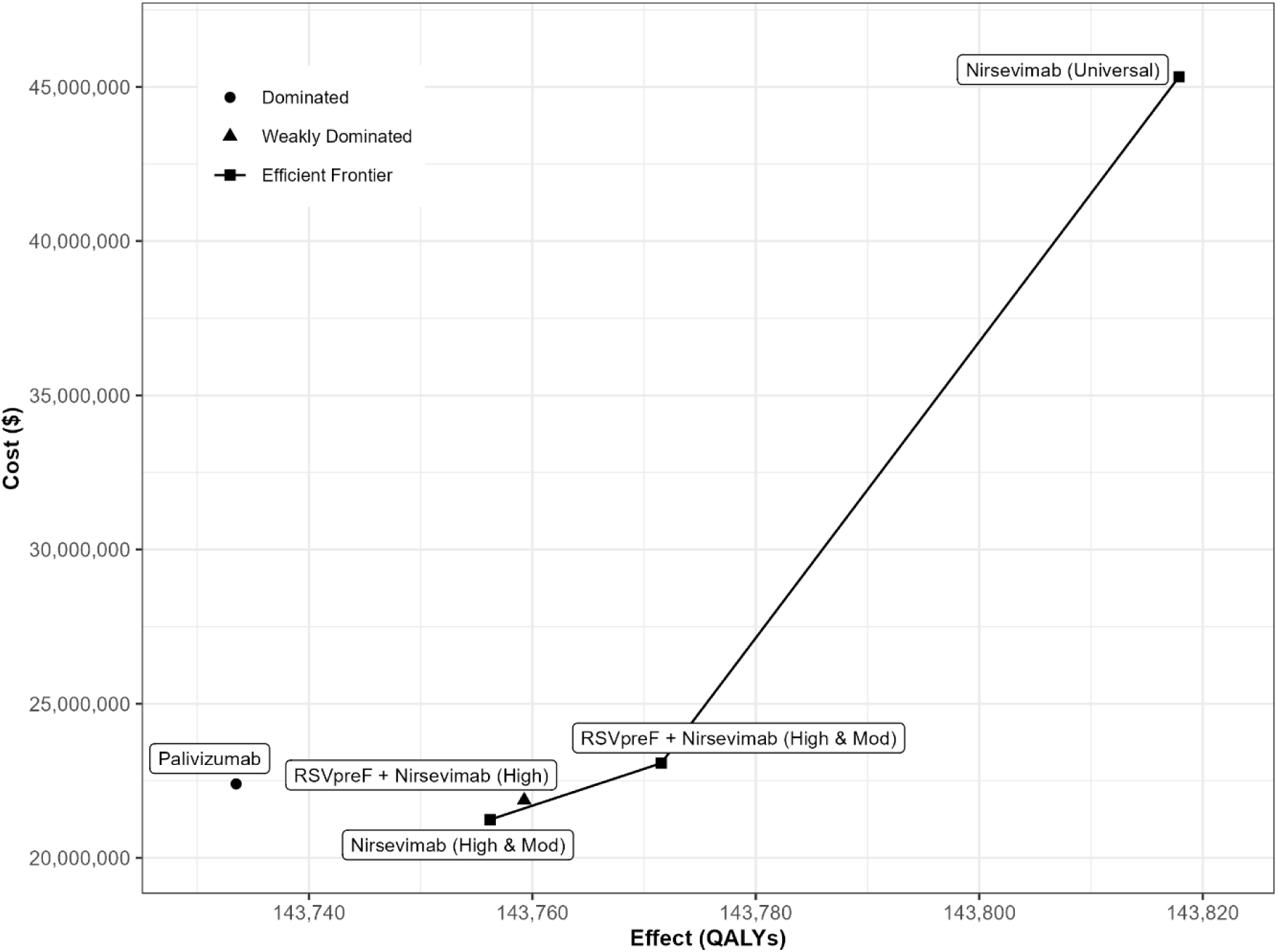
Results of the sequential cost-effectiveness analysis comparing all RSV prevention strategies to each other and to the standard of care, from the health system perspective.

**Table 4.** Cost-effectiveness results for alternative RSV immunization strategies. Table 4A presents the sequential analysis comparing non-dominated strategies stepwise. Table 4B shows the pairwise cost-effectiveness of each strategy relative to the historical standard of care.

4A. Sequential cost-effectiveness analysis
| Strategies | Incremental Cost | Incremental QALY | ICER |
| --- | --- | --- | --- |
| Nirsevimab (High & Mod) | - | - | - |
| RSVpreF + Nirsevimab (High & Mod) | \$1,828,392 | 15 | \$119,418 /QALY |
| Nirsevimab (All-infant) | \$22,256,546 | 46 | \$480,253/QALY |

4B. Pairwise cost-effectiveness results
| Strategies | Incremental Cost | Incremental QALY | ICER |
| --- | --- | --- | --- |
| Palivizumab (High) | - | - | - |
| Nirsevimab (High & Mod) | -\$1,158,224 | 23 | Dominant |
| RSVpreF + Nirsevimab (High) | -\$531,712 | 26 | Dominant |
| RSVpreF + Nirsevimab (High & Mod) | \$670,168 | 38 | \$ 17,611/QALY |
| Nirsevimab (All-infant) | \$22,926,715 | 84 | \$ 271,651/QALY |

In the sequential analysis (Table 4A), nirsevimab for high- and moderate-risk infants emerged as the most cost-effective non-dominated strategy, improving health outcomes while reducing overall costs relative to the next-best alternative. Replacing palivizumab for high-risk infants with nirsevimab for both high- and moderate-risk groups generated approximately $1.2 million in savings and yielded 23 aditional QALYs. Expanding coverage further to include maternal RSVpreF vaccination plus nirsevimab for high- and moderate-risk infants increased health gains but required additional investment (approximately $1.8 million for 15 QALYs gained), resulting in an ICER of about $119,000 per QALY. Although this combined strategy remains on the efficient frontier, its ICER exceeds the conventional threshold. When compared directly with the historical standard of care, however, the same strategy was cost-effective (ICER ≈ $18,000 per QALY; Table 4B). Regardless of the comparative framework adopted, nirsevimab for all infants remained not cost-effective under current assumptions.

### 3.3. Sensitivity analysis

We conducted additional analyses to assess the robustness of our base-case results. The one-way sensitivity analysis examined how uncertainty in key parameters influences the expected net monetary benefit (NMB) across all strategies. To improve readability, the all-infant nirsevimab strategy (which consistently had much lower NMB values and never became more efficient than any other option across the entire parameter range) was excluded from the main figure. Figure 5 illustrates how NMB changes as each parameter is varied across its plausible range while holding all others constant. Across all strategies, the most influential parameters were the unit costs and efficacies of RSVpreF and nirsevimab. Other important factors included the efficacy of palivizumab and the per-day costs of pediatric ward and ICU care. The complete version of this analysis, including the all-infant nirsevimab strategy, is presented in Supplementary Figure S3.

**Figure 5.**
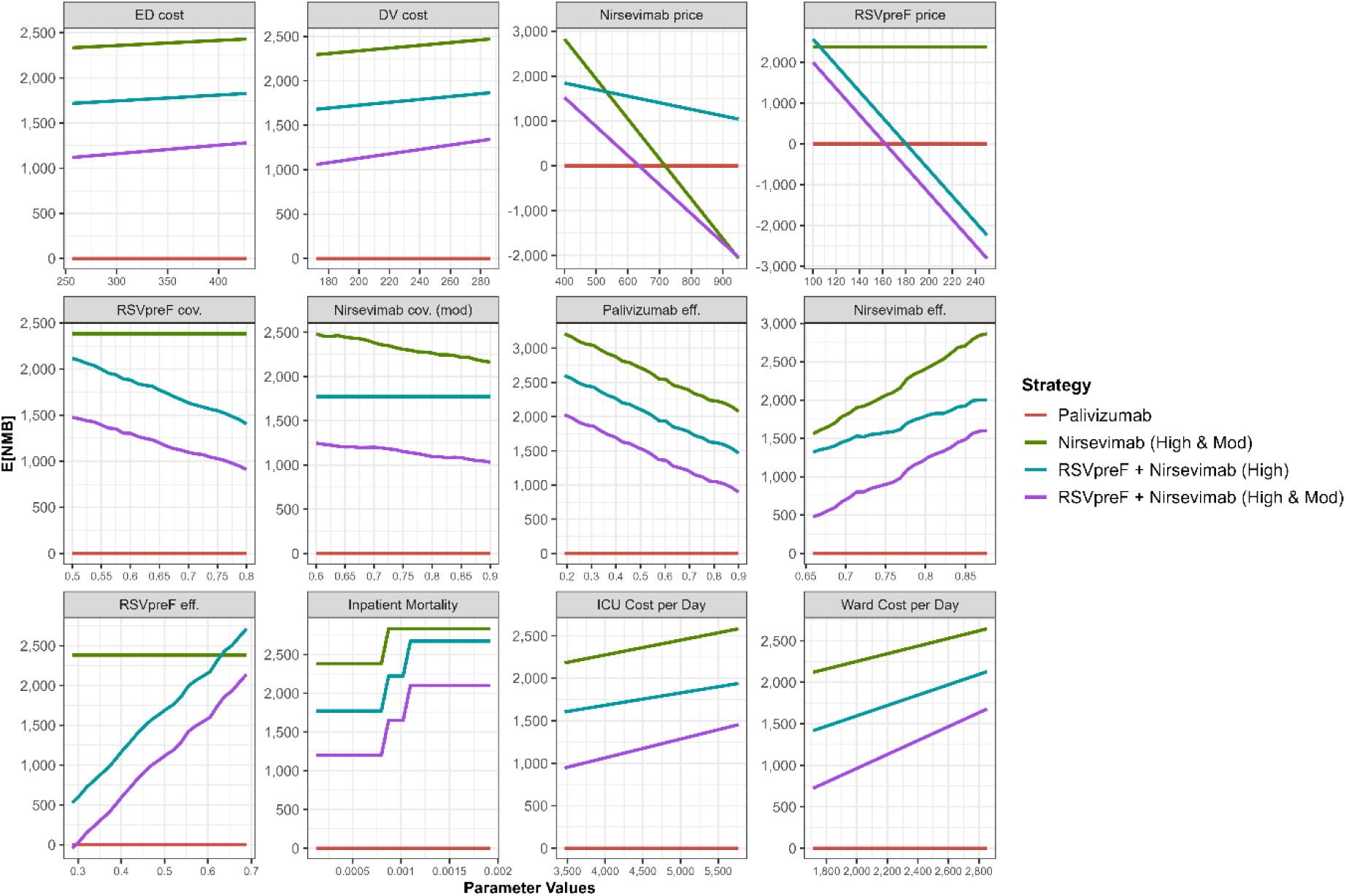
One-way sensitivity analysis showing the expected Net Monetary Benefit (E[NMB]). Each parameter was varied independently across its plausible range while holding all others constant. The outcome represents E[NMB], expressed in thousands of Canadian dollars.

Additionally, we performed a two-way sensitivity analysis to explore the simultaneous variation of RSVpreF and nirsevimab unit costs. Figure 6 indicates the immunization strategy yielding the highest NMB across a range of plausible price combinations (nirsevimab: $0–$1000 per dose; RSVpreF: $0–$300 per dose). For most price scenarios, the highest NMB was achieved by either (a) maternal RSV vaccination for in-season births combined with nirsevimab for high-risk children, or (b) nirsevimab for both high- and moderate-risk children. The fully combined strategy (maternal vaccination plus nirsevimab for both high- and moderate-risk children) generated the highest NMB within only a narrow price range. Notably, for all-infant nirsevimab to yield the highest NMB, its price would need to drop below approximately $110 per dose, well below current estimates.

**Figure 6.**
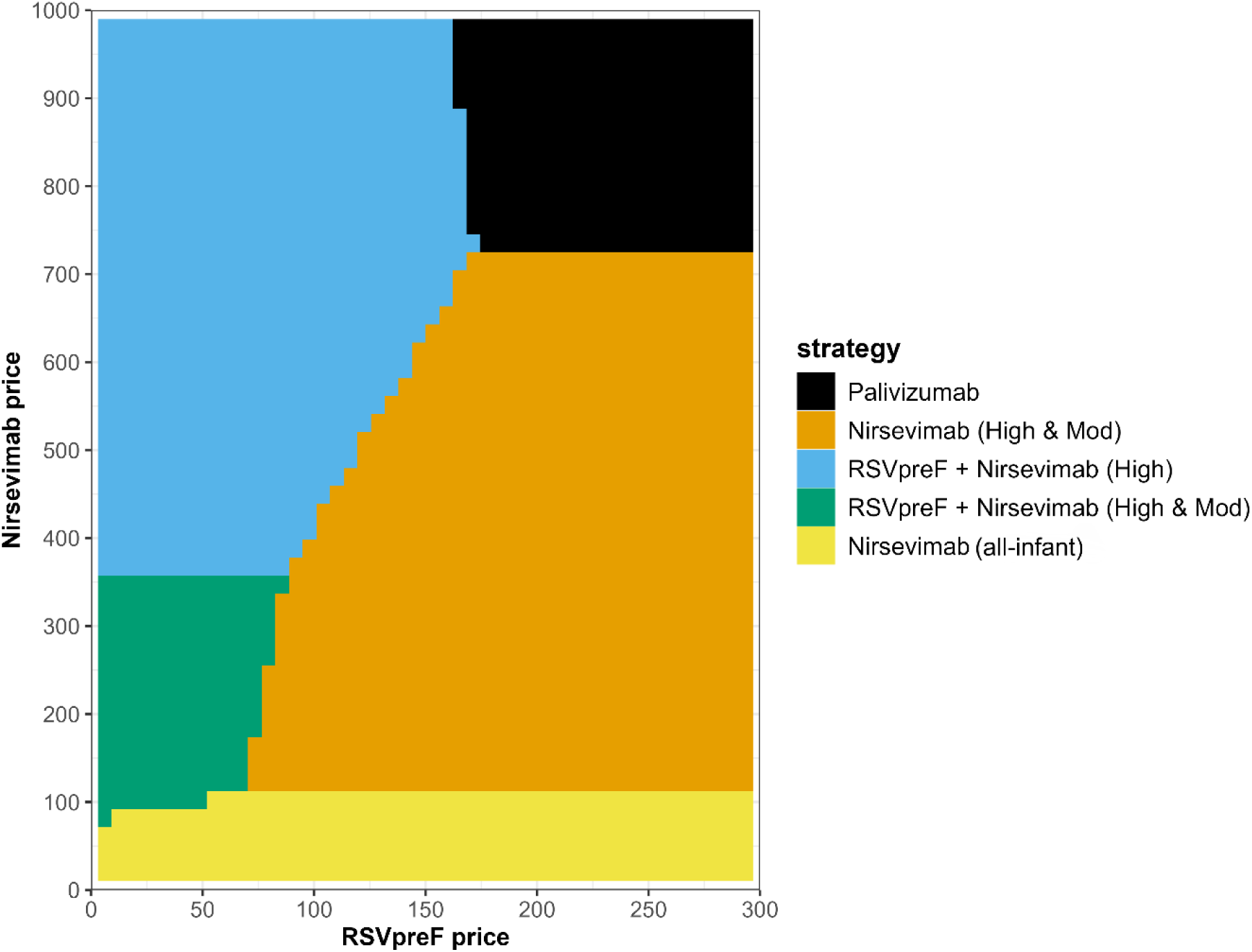
Two-way sensitivity analysis varying the unit prices of nirsevimab and RSVpreF. Each region on the plot indicates the strategy yielding the highest NMB given a specific combination of nirsevimab and RSVpreF prices, including the historical standard of care for reference.

## Discussion

Our evaluation provides compelling evidence that targeted RSV immunization strategies offer significantly greater economic value than both the historical palivizumab program and an all-infant nirsevimab prophylaxis approach. Specifically, replacing palivizumab for high-risk children, with nirsevimab for both high- and moderate-risk children led to improved health outcomes for a broader group of children while reducing overall healthcare expenditures. The combined maternal RSVpreF plus nirsevimab (high & moderate) strategy yielded additional health gains but was not cost-effective incrementally (ICER ≈ $119,000/QALY vs nirsevimab (high & moderate)). However, when evaluated directly against the historical standard of care (Palivizumab was the immunization program in place at the time the study was conducted), this combined approach remained cost-effective (ICER ≈ $18,000/QALY), suggesting value if introduced as a program expansion. In contrast, nirsevimab for all infants was not cost-effective under base-case assumptions.

These findings partially align with the current direction of national immunization policy in Canada. NACI has recently recommended a staged expansion of RSV immunization programs, beginning with high-risk infants and working toward making RSV immunization available to all infants [6]. This approach reflects constraints related to supply, cost-effectiveness, and fiscal sustainability. While our analysis supports a targeted and phased rollout, it identifies a different optimal pathway for program expansion. Specifically, the combined maternal RSVpreF plus nirsevimab (high & moderate) strategy provides substantially greater economic value than a universal all-infant nirsevimab program at current prices, delivering more health benefit per dollar spent. In addition to offering greater economic efficiency, the combined maternal RSVpreF plus nirsevimab (high & moderate) strategy would require a far smaller budget increase than a universal all-infant nirsevimab program. In contrast, universal nirsevimab would become economically attractive only if the unit price were to decline substantially.

Our study also builds on prior economic evaluations of RSV immunization in Canada and internationally. Shoukat et al. employed a similar simulation-based approach based on data from Ontario, and concluded that all-infant nirsevimab prophylaxis would only be cost-effective if the unit price fell to roughly $290 per dose or lower [7]. They found that a strategy combining maternal RSV vaccination with targeted nirsevimab for high-risk infants achieved substantial reductions in infant mortality (∼76–85%) and was more economically efficient than all-infant immunization.

These findings parallel our own, with both studies highlighting that targeted strategies can provide nearly equivalent health benefits at a lower cost. Similarly, Shin et al. used a static decision tree model to assess an all-infant nirsevimab program across Canadian infants, reporting that such a strategy would be cost-effective if the price per dose remained below approximately $550 under a $50,000/QALY willingness-to-pay threshold [21]. Their model projected large reductions in RSV-related healthcare events and direct costs; however, the inputs for RSV mortality and intensive care utilization were higher than those observed at the population level in BC. Another national-level analysis by Gebretekle et al. examined multiple immunization strategies across the first RSV season and found that neither an all-infant nirsevimab approach nor a year-round maternal RSVpreF program were cost-effective at list prices [8]. Their model identified seasonal nirsevimab administration with “catch-up” dosing for infants born just before the RSV season as the most economically favorable strategy, especially when focused on high- and moderate-risk infants. Notably, the list prices assumed for nirsevimab and RSVpreF in their analysis were nearly double the prices used in our study, which likely contributed to their more conservative cost-effectiveness conclusions for broader strategies. Their findings are particularly notable in remote and high-cost settings, where ICERs were as low as $5,700 per QALY. These Canadian studies, together with ours, support the conclusion that risk-based targeting of RSV immunoprophylaxis maximizes value for money.

Although these studies assumed higher mortality rates among hospitalized infants, in-hospital case fatality attributable to RSV remains very low in high-income countries [1,22], consistent with BC’s recent experience of approximately one RSV-associated infant death every 2–3 years [10]. This low mortality likely reflects the impact of longstanding preventive programs (e.g., palivizumab for high-risk infants), advanced pediatric care, and potentially lower viral transmission due to demographic or geographic factors [9]. As such, models assuming uniformly high mortality among hospitalized infants may overestimate the number of deaths averted by a transition to expanded immunization programs. To avoid overstating the potential life-saving impact and cost-effectiveness of RSV interventions, we applied a conservative baseline mortality assumption.

International analyses also reinforce the prioritization of targeted RSV immunization and highlight the influence of health system context. In the United States, where both nirsevimab and RSVpreF have recently been authorized, Averin et al. evaluated cost-effectiveness under multiple program designs. Their analysis showed that a year-round maternal vaccination strategy would prevent about 12% of medically attended RSV cases yielding an ICER of $90,000 USD per QALY [23]. By contrast, a seasonal maternal program—targeting only pregnancies due during the RSV season— was found to be cost-saving from a societal perspective despite preventing fewer total cases. These results support a seasonal maternal immunization strategy, emphasizing the importance of targeting protection to periods of peak vulnerability. In Europe, Getaneh et al. assessed RSV prevention strategies in six countries and found that seasonal all-infant immunization with a catch-up dose was generally the most cost-effective strategy when the prophylaxis price was low (€50 per dose) [24]. While maternal vaccination had lower impact on infant outcomes, it became the preferred strategy in specific contexts (e.g., Veneto, Italy) or under more generous willingness-to-pay thresholds.

From a methodological standpoint, our study introduces several important innovations relative to earlier work. First, we addressed a key limitation in prior Canadian models by incorporating both age and seasonality into RSV risk estimation. Risk of infection and severity is highest during the first months of life and peaks in winter, but earlier models did not fully capture this interaction. Previous studies also did not consider that the severity of RSV illnesses is not uniform across children’s groups, and that children with co-morbidities require significantly longer hospitalizations stays and a higher proportion of them required ICU admissions [9]. By implementing a discrete-event simulation with monthly time steps and stratifying children by birth month, we were able to apply joint age–season risk profiles when estimating probabilities of hospitalization and ICU admission. This modeling structure allowed for a more precise evaluation of interventions such as in-season maternal vaccination, which is most effective for children born just before or during the RSV season. It also adjusts for the potential additional health gains of immunization in children’s groups who are most vulnerable. Second, by following children over two full RSV seasons, our model captures benefits that extend into the second year of life. Third, we used actual RSV outcome data representing the entire BC population, which provide population-level RSV hospitalization and ICU risks. This contrasts with previous studies that relied on tertiary care hospital data drawn from diverse sources and regions, which may not reflect consistent patterns [7,8]. Additionally, the use of current healthcare cost data, including estimates of pediatric ward and ICU costs provide more contemporary estimates of costs, which have increased substantially over the past decade. By contrast, many prior analyses relied on cost estimates from 2017, potentially underestimating the economic burden of severe RSV illness and thus the potential savings from prevention [7,8].

Our sensitivity analyses further bolster the robustness of our findings. One-way analyses of key parameters—including vaccine efficacy, hospitalization costs, and unit prices—showed that the qualitative result remained unchanged: targeted strategies consistently outperformed both the historical standard of care and all-infant immunization. Even under conservative assumptions, the maternal plus nirsevimab (high & moderate) strategy yielded a higher NMB compared to the historical standard of care. Our two-way sensitivity analysis on intervention prices was particularly informative, identifying the price combinations at which the preferred strategy would shift. The cost threshold for nirsevimab for all-infant programs to be considered cost-effective is consistent with findings from Gebretekle and Shoukat. This alignment across studies enhances the credibility of our conclusions and affirms the central role of price and risk stratification in determining the value of RSV immunization strategies.

Nevertheless, our analysis has some limitations. First, we assumed a simplified waning pattern for immunity: constant protection for five months post-immunization, followed by linear decline to zero by ten months. While this assumption aligns with clinical trial follow-up durations, real-world effectiveness may differ. If protection extends beyond ten months, the cost-effectiveness of interventions would be underestimated in our model. Conversely, if immunity wanes more rapidly, particularly in specific subgroups, our results may overstate benefits. More empirical data on real-world durability of RSVpreF and nirsevimab will be essential to refine future models. Second, we hypothesized a higher uptake rate for infant nirsevimab prophylaxis than for maternal RSVpreF vaccination, whereas emerging Canadian data indicate that families may prefer maternal immunization [25]. This preference could affect the relative cost-effectiveness of combined maternal-plus-infant immunization strategies.

In addition, our model used a static cohort design and did not account for indirect effects on RSV transmission. While static models assume each child’s infection risk is independent of others, dynamic models allow for herd effects, age shifts in incidence, and broader population-level impacts. Previous work has shown that dynamic transmission models often predict different patterns of benefit. For instance, Li et al. observed that infant-focused immunization may delay, rather than prevent, some RSV infections, shifting disease burden to older children [26]. Similarly, Hodgson et al. used a dynamic transmission model in the UK and found that while large-scale RSV immunization programs can reduce overall virus circulation, they may also lead to indirect effects— such as changes in the age distribution of infections or transmission patterns—that must be carefully considered in policy decisions [27]. Our static model likely overestimates the total number of infections averted, but severe outcomes in children (hospitalizations and deaths) are more robust to model choice. Since our analysis focuses on cost and QALYs from the healthcare payer perspective over a two-year child horizon, we expect the core conclusions about cost-effectiveness of targeted strategies to remain valid.

Finally, we adopted a health system perspective, excluding societal costs such as caregiver productivity losses, travel expenses, and the psychosocial impact on families. This is a conservative approach that likely underestimates the full value of immunization. RSV-related hospitalizations impose substantial non-medical costs, particularly in rural or remote regions where medical evacuations are frequent and costly. Including these broader costs—as done in Gebretekle et al. [8] and Getaneh et al. [24]—would likely make prophylactic interventions appear even more cost-effective. Additionally, our analysis did not explicitly model cost-effectiveness in remote or isolated settings. In such contexts, the substantially higher transport and hospitalization costs associated with severe RSV cases could make broader prophylaxis approaches (such as a universal infant Nirsevimab program) more economically favorable than in the provincial base-case scenario. Our QALY estimates also captured only the health-related utility loss in children, not the quality-of-life burden on caregivers or potential long-term morbidity. Including these outcomes in future models would provide a more comprehensive assessment of value.

## 4. Conclusion

This study demonstrates that replacing the palivizumab program, which was the standard of care at the time the study was conducted, with nirsevimab for high- and moderate-risk children is both cost-saving and more effective in preventing RSV-related illness. Adding seasonal maternal RSVpreF vaccination to this strategy yields additional health benefits and remains cost-effective when compared with the historical standard of care, though it exceeds the willingness-to-pay threshold in the sequential analysis. This suggests it represents a valuable program expansion option if resources permit. In contrast, all-infant immunization with nirsevimab is not economically justified without a substantial price reduction. These findings support a phased approach to RSV immunization, starting with the replacement of palivizumab with nirsevimab and extending its use to moderate-risk infants, followed by maternal RSVpreF vaccination to protect low-risk infants as costs decrease. This approach aligns with national recommendations and offers a pathway to optimize both health outcomes and healthcare resource use under current pricing and supply conditions for RSV immunization products.

## Data Availability

Access to data provided by the Data Stewards is subject to approval but can be requested for research projects through the Data Stewards or their designated service providers.

https://www2.gov.bc.ca/gov/content/health/health-forms/online-services

https://www2.gov.bc.ca/gov/content/health/conducting-health-research-evaluation/data-access-health-data-central

## CRediT authorship contribution statement

All authors attest they meet the ICMJE criteria for authorship. Contributions were as follows: **JT:** Conceptualization, Methodology, Software, Formal analysis, Data Curation, Writing - Original Draft, Writing - Review & Editing, Supervision. **MVP:** Software, Formal analysis, Data Curation, Writing - Review & Editing. **AW:** Data Curation, Writing - Review & Editing. **MC:** Formal analysis, Writing - Review & Editing**. JMHW:** Formal analysis, Writing - Review & Editing**. JP:** Formal analysis, Writing - Review & Editing**. LS:** Formal analysis, Writing - Review & Editing**. Jia Hu** Formal analysis, Writing - Review & Editing**. DS:** Formal analysis, Writing - Review & Editing**. PML:** Conceptualization, Formal analysis, Writing - Review & Editing, Supervision**. HS:** Conceptualization, Methodology, Formal analysis, Data Curation, Writing - Review & Editing, Supervision.

## Data availability

Access to data provided by the Data Stewards is subject to approval but can be requested for research projects through the Data Stewards or their designated service providers. The following data sets were used in this study: Client Roster (CR), Discharge Abstracts Database (DAD), BC Vital Statistics (VS) [12–14]. All inferences, opinions, and conclusions drawn in this publication are those of the author(s), and do not reflect the opinions or policies of the Data Steward(s).

