## supplementary material for "Cost-Effectiveness of Infant and Maternal RSV Immunization Strategies, in British Columbia, Canada"

### 1. RSV Hospitalization Risk Model

RSV hospitalization risk in infancy is highest at younger ages and during peak winter months. Figure S1 illustrates the observed RSV hospitalization rates by infant age and month of infection, based on epidemiological data specific to British Columbia [1–3].

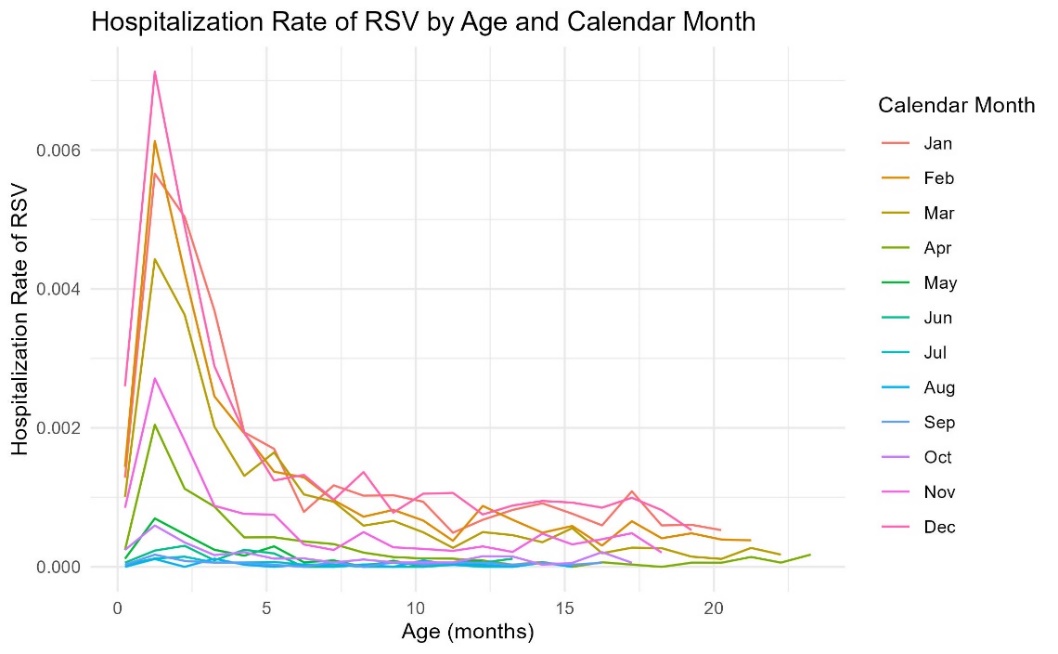

Figure S1 RSV hospitalization rate (per live birth) by infant age and month of infection (BC data).

We used a logistic regression model to estimate an infant’s monthly probability of RSV hospitalization as a function of age, PVZ prophylaxis status, risk group, and seasonality. The model included non-linear terms for age, a binary variable for prophylaxis status (PVZ), and indicator variables for risk category (moderate-risk, high-risk) and calendar month of infection (Month). The form of the logistic model is given by the equation below:

$${Logit(prob}_{hospitalization})= \beta_{0}+\beta_{1}\left( \frac{1}{Age} \right)+ \beta_{2}Age+ \beta_{3}Age^{2}+\beta_{4}Age^{3}+PVZ+Risk+Month+e$$

Equation S1 Logistic regression model estimating the monthly probability of RSV hospitalization among infants as a function of age, prophylaxis status (PVZ), risk group, and calendar month.

The estimated model coefficients are presented in Table S1.

Table S1 Estimated Coefficient of the model for probability of hospitalization.

| Variable | Estimated Coefficient | Confidence Interval |
| --- | --- | --- |
| (Intercept) | -7.77 | (-8.25 – -7.29) |
| I(1/Age) | -0.63 | (-0.68 – -0.58) |
| Age | -0.76 | (-0.82 – -0.70) |
| I(Age2) | 0.056 | (0.049 – 0.062) |
| I(Age3) | -0.0014 | (-0.0016 – -0.0012) |
| PVZ | -0.56 | (-1.02 – -0.09) |
| Risk (Moderate Risk) | 0.85 | (0.78 – 0.92) |
| Risk (High Risk) | 1.79 | (1.37 – 2.21) |
| Calendar month 1 | 3.96 | (3.50 – 4.43) |
| Calendar month 2 | 3.82 | (3.36 – 4.29) |
| Calendar month 3 | 3.56 | (3.09 – 4.02) |
| Calendar month 4 | 2.46 | (1.98 – 2.94) |
| Calendar month 5 | 1.56 | (1.05 – 2.07) |
| Calendar month 6 | 1.04 | (0.49 – 1.58) |
| Calendar month 7 | 0.20 | (-0.43 – 0.82) |
| Calendar month 9 | 0.49 | (-0.10 – 1.07) |
| Calendar month 10 | 1.61 | (1.11 – 2.11) |
| Calendar month 11 | 3.04 | (2.56 – 3.51) |
| Calendar month 12 | 4.05 | (3.59 – 4.52) |

### 2. Vaccine Efficacy and Coverage Assumptions

Table S2 summarizes the vaccine efficacy and coverage inputs used for each strategy.

Table S2 Vaccine coverage and efficacy inputs.

| Parameter | Base value | Range | Reference |
| --- | --- | --- | --- |
| Uptake (coverage) | | | |
| RSVpreF maternal vaccine coverage (% of eligible pregnancies) | 65% | (50-80) | [4] |
| Nirsevimab coverage (% of infants at moderate/low risk) | 70% | (60-90) | [5] |
| Palivizumab coverage for high-risk children in first year | 100% |  | BC internal data |
| Palivizumab coverage for high-risk children in second year | 16% | (15 – 17) | BC internal data |
| Palivizumab efficacy (1 month post-immunization) | | | |
| Efficacy against medically attended RSV | 70 % | (19–90) | [5] |
| Efficacy against RSV hospitalization | 82 % | (29–96) | [5] |
| Efficacy against RSV ICU admission | 82 % | (29–96) | [5] |
| RSVpreF efficacy (0–5 months post-immunization) | | | |
| Efficacy against medically attended RSV | 52.5% | (28.7–68.9) | [6] |
| Efficacy against RSV hospitalization | 56.4% | (5.2–81.5) | [6] |
| Efficacy against RSV ICU admission | 70.9% | (44.5–85.9) | [6] |
| Nirsevimab efficacy (0–5 months post-immunization) | | | |
| Efficacy against medically attended RSV | 79·5% | (65·9–87·7) | [7] |
| Efficacy against RSV hospitalization | 77·3% | (50·3–89·7) | [7] |
| Efficacy against RSV ICU admission | 86·0% | (62·5–94·8) | [7] |

Figure S2 illustrates our assumptions regarding vaccine efficacy waning over time. We modeled the effectiveness of both RSVpreF and nirsevimab as remaining constant for the first five months following immunization, after which efficacy declines linearly to zero by the tenth month.

Figure S2 Efficacy waning over time for (A) nirsevimab and (B) maternal RSVpreF.

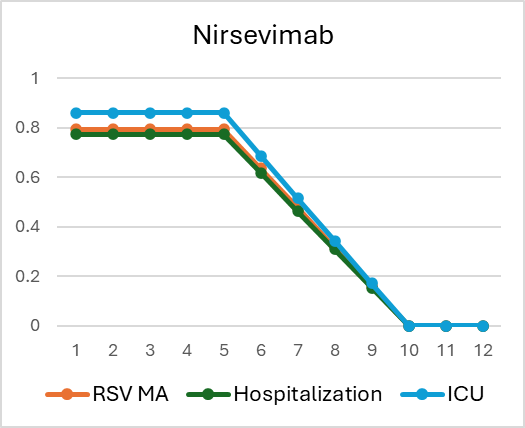

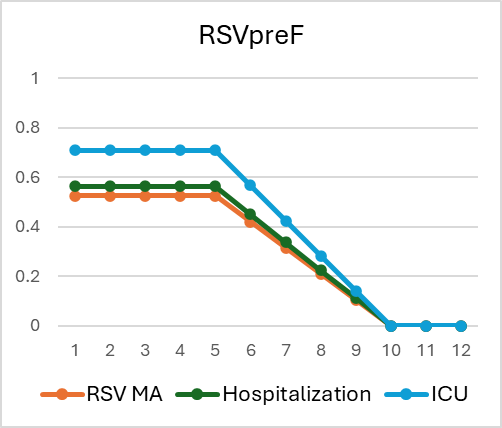

**(A)**

**(B)**

Nirsevimab

|  | RSV MA | Hospitalization | ICU |
| --- | --- | --- | --- |
| 1 | 0.795 | 0.773 | 0.86 |
| 2 | 0.795 | 0.773 | 0.86 |
| 3 | 0.795 | 0.773 | 0.86 |
| 4 | 0.795 | 0.773 | 0.86 |
| 5 | 0.795 | 0.773 | 0.86 |
| 6 | 0.636 | 0.6184 | 0.688 |
| 7 | 0.477 | 0.4638 | 0.516 |
| 8 | 0.318 | 0.3092 | 0.344 |
| 9 | 0.159 | 0.1546 | 0.172 |
| 10 | 0 | 0 | 0 |
| 11 | 0 | 0 | 0 |
| 12 | 0 | 0 | 0 |

RSVpreF

|  | RSV MA | Hospitalization | ICU |
| --- | --- | --- | --- |
| 1 | 0.525 | 0.564 | 0.709 |
| 2 | 0.525 | 0.564 | 0.709 |
| 3 | 0.525 | 0.564 | 0.709 |
| 4 | 0.525 | 0.564 | 0.709 |
| 5 | 0.525 | 0.564 | 0.709 |
| 6 | 0.42 | 0.4512 | 0.5672 |
| 7 | 0.315 | 0.3384 | 0.4254 |
| 8 | 0.21 | 0.2256 | 0.2836 |
| 9 | 0.105 | 0.1128 | 0.1418 |
| 10 | 0 | 0 | 0 |
| 11 | 0 | 0 | 0 |
| 12 | 0 | 0 | 0 |

### 3. Additional Sensitivity Analysis

We conducted additional one-way sensitivity analyses to assess the robustness of our findings across all immunization strategies, including the universal Nirsevimab option. For each key parameter, the expected net monetary benefit (E[NMB]) was recalculated while varying that parameter across its plausible range and holding all others constant.

Figure S3 presents the results for all strategies, demonstrating how E[NMB] changes with each parameter. The universal Nirsevimab strategy consistently exhibited substantially lower E[NMB] values than other options and did not become more efficient than any other strategy across the entire parameter range.

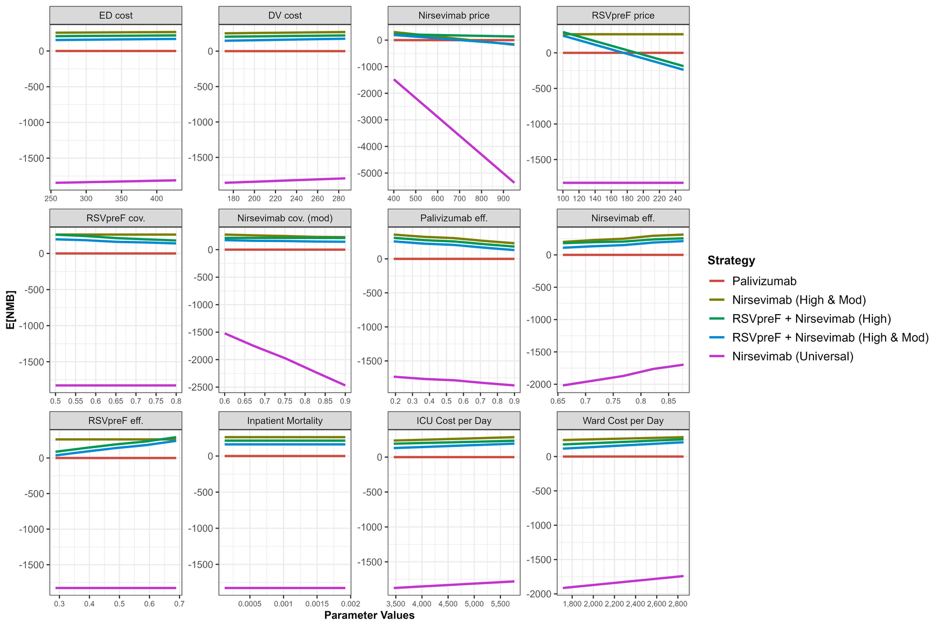

Figure S3 One-way sensitivity analysis showing the expected Net Monetary Benefit (E[NMB]) for the standard of care and all immunization strategies, including the universal Nirsevimab strategy. Each parameter was varied independently across its plausible range while holding all others constant. The outcome represents E[NMB], expressed in thousands of Canadian dollars.

For comparison, Figure S4 displays the tornado diagram summarizing the influence of individual parameters on the combined maternal RSVpreF plus infant nirsevimab (high- and moderate-risk) strategy relative to the historical standard of care. Similar to the main findings, the unit costs and efficacies of RSVpreF and nirsevimab remained the most influential determinants of cost-effectiveness.

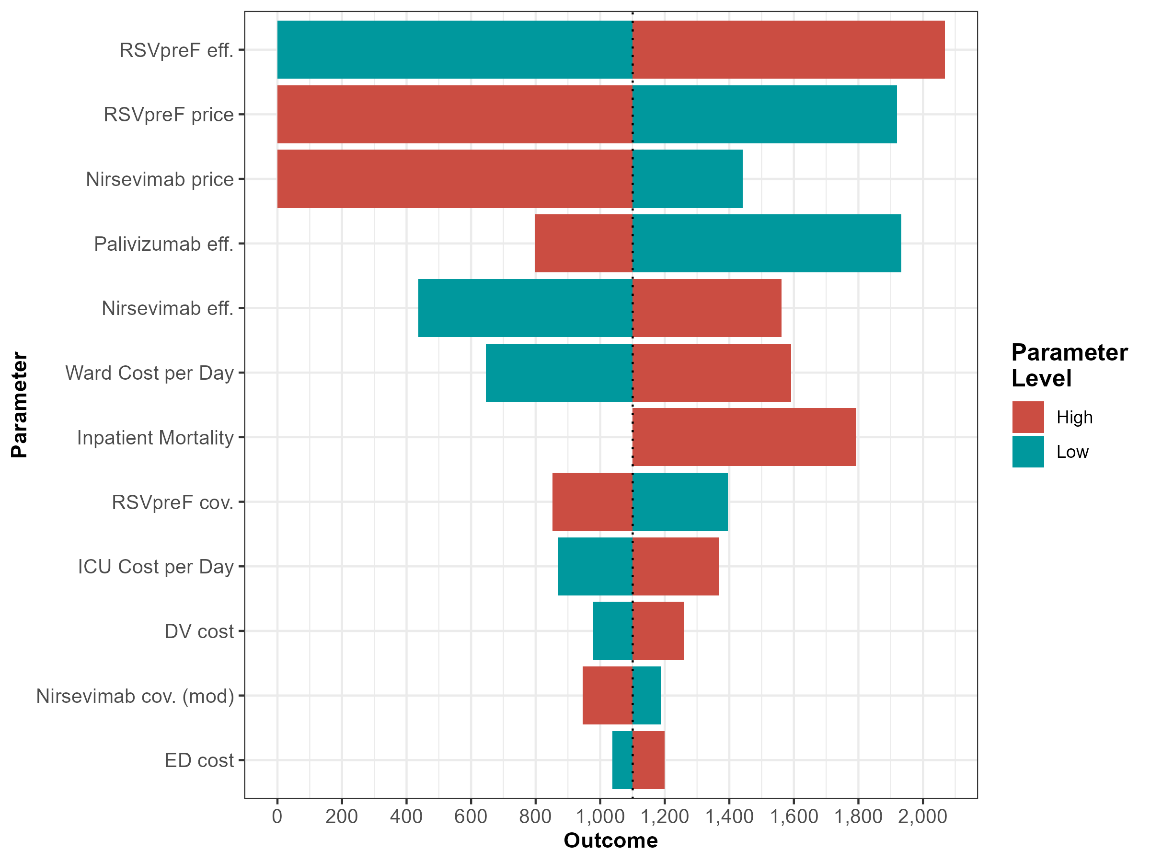

Figure S4 Tornado diagram summarizing the one-way sensitivity analysis for the combined maternal RSVpreF and infant nirsevimab (high- and moderate-risk) strategy relative to the historical standard of care. Bars show the change in NMB as each parameter is varied across its plausible range, with longer bars indicating greater influence on model results
